# Virtual brain grafting: Enabling whole brain parcellation in the presence of large lesions

**DOI:** 10.1101/2020.09.30.20204701

**Authors:** Ahmed M. Radwan, Louise Emsell, Jeroen Blommaert, Andrey Zhylka, Silvia Kovacs, Tom Theys, Nico Sollmann, Patrick Dupont, Stefan Sunaert

## Abstract

Brain atlases and templates are at the heart of neuroimaging analyses, for which they facilitate multimodal registration, enable group comparisons and provide anatomical reference. However, as atlas-based approaches rely on correspondence mapping between images they perform poorly in the presence of structural pathology. Whilst several strategies exist to overcome this problem, their performance is often dependent on the type, size and homogeneity of any lesions present. We therefore propose a new solution, referred to as Virtual Brain Grafting (VBG), which is a fully-automated, open-source workflow to reliably parcellate MR images in the presence of a broad spectrum of focal brain pathologies, including large, bilateral, intra- and extra-axial, heterogeneous lesions with and without mass effect.

The core of the VBG approach is the generation of a lesion-free T1-weighted input image which enables further image processing operations that would otherwise fail. Here we validated our solution based on Freesurfer recon-all parcellation in a group of 10 patients with heterogeneous gliomatous lesions, and a realistic synthetic cohort of glioma patients (n=100) derived from healthy control data and patient data.

We demonstrate that VBG outperforms a non-VBG approach assessed qualitatively by expert neuroradiologists and Mann-Whitney U tests to compare corresponding parcellations (real patients U(6,6) = 33, z = 2.738, *P* < .010, synthetic patients U(48,48) = 2076, z = 7.336, *P* < .001). Results were also quantitatively evaluated by comparing mean dice scores from the synthetic patients using one-way ANOVA (unilateral VBG = 0.894, bilateral VBG = 0.903, and non-VBG = 0.617, *P* < .001). Additionally, we used linear regression to show the influence of lesion volume, lesion overlap with, and distance from the Freesurfer volumes of interest, on labelling accuracy.

VBG may benefit the neuroimaging community by enabling automated state-of-the-art MRI analyses in clinical populations, for example by providing input data for automated solutions for fiber tractography or resting-state fMRI analyses that could also be used in the clinic. To fully maximize its availability, VBG is provided as open software under a Mozilla 2.0 license (https://github.com/KUL-Radneuron/KUL_VBG).

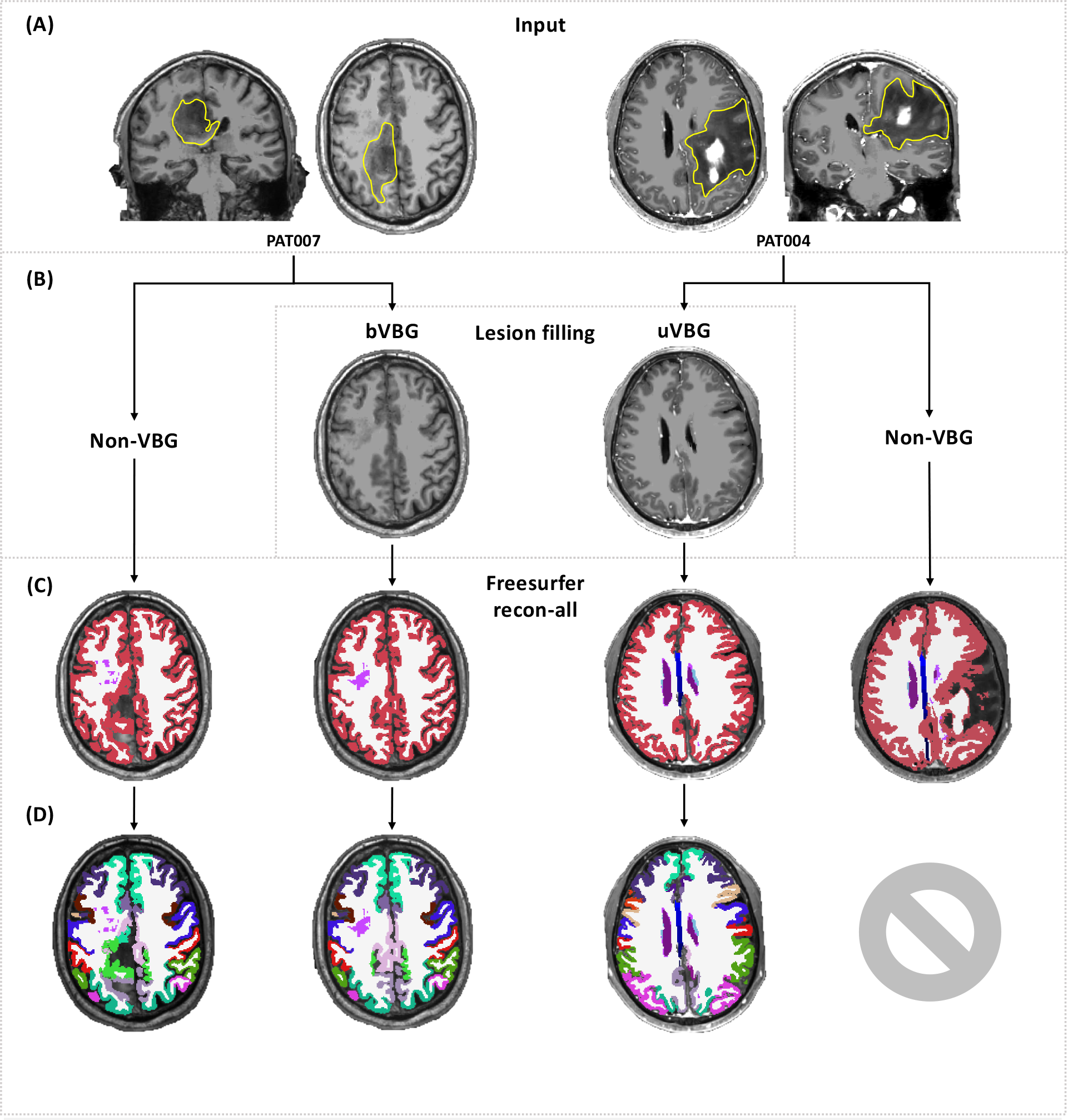
Graphical abstract: (A) shows T1 images from two patients with gliomatous lesions. VBG is a lesion replacement/filling workflow with one approach for unilateral lesions (uVBG) and another for bilateral lesions (bVBG). (B) shows the recon-all approach selected, (C) & (D) show the output, tissue segmentations (C) and whole brain parcellations (D). If VBG is not used (non-VBG) recon-all may finish with some errors in the parcellations (left) or fail to generate a parcellation entirely (right). However, using either VBG method allows recon-all to complete where it had previously failed and also improves parcellation quality.

## 1. Introduction

Brain mapping is the process of spatially representing known brain structures, e.g. anatomical regions, or functional areas on a coordinate system or space. Attempts to create a map of the human brain predate the 20^th^ century (Moore, 2011; Standring, 2016), initially existing mostly in anatomical and medical textbooks as sketches of ex-vivo specimens. The earliest modern detailed cytoarchitectonic representation of the human brain is the Brodmann atlas, published in a monograph in 1909 and remains widely used in the field today (Mandal et al., 2012). Thanks to Talairach and Tournoux (Talairach and Tournoux, 1988) and similar coordinate systems as well as other advances in medical imaging and computer science, brain atlases are now widely used in neuroimaging and neurosurgery for structural brain mapping (Desikan et al., 2006; Dickie et al., 2017; Frazier et al., 2005; Goldstein et al., 2007; Makris et al., 2006; Nowinski, 2016; Tzourio-Mazoyer et al., 2002). Atlas-based brain mapping can be achieved accurately and non-invasively for individual subjects or even groups using brain images acquired from a magnetic resonance imaging (MRI) scanner. Many atlases are available non-commercially as digital image files, representing different brain structures using specific image intensity values (Dickie et al., 2017).

Typically, brain atlas images are defined in the space of a template or reference brain image, which may be a single subject image or an averaged image from a specific cohort, e.g. Montreal Neurological Institute 152 (MNI152) brain template (Mazziotta et al., 1995). Structural brain mapping using an atlas requires it first to be spatially matched to the brain image being mapped, or vice versa. This can be achieved by image registration, a process that maximizes spatial similarity between source and target images, also known as normalization, when a standard brain image such as MNI152 is the target. Many strategies exist, including rigid-body and affine registration, and non-affine approaches such as symmetric diffeomorphic registration (Avants et al., 2008). If successful, the resulting transforms and/or warps can be used to bring the atlas to the patient’s brain or, vice versa, achieving a basic brain mapping, which may be efficient but lacking in tissue and subject specificity. More advanced pipelines e.g. Freesurfer (Fischl, 2012) recon-all (FreeSurferWiki, 2020a) leverage several steps e.g. brain extraction, tissue segmentation and surface-based registration to maximize tissue and subject specificity.

Structural brain mapping, also called segmentation or parcellation, can normally be automated, and is available through several programs such as FSL (Jenkinson et al., 2012), SPM (Ashburner et al., 2006), ANTs (Avants et al., 2011), Freesurfer (Fischl, 2012), AFNI (Cox, 1996) and others (GitHub, 2020; Hanke and Halchenko, 2011; Jahn, 2020; NeuroDebian, 2020; NITRC, 2020). However, as this process typically relies on image contrast between tissues and prior anatomical knowledge, most methods are optimized for anatomically normal or close to normal brains without major morphologic aberrations. Consequently, brain parcellation remains problematic in the presence of structural brain pathology (Ledig et al., 2015; Nachev et al., 2008; Solodkin et al., 2010; Weiller et al., 1995). It is important to realize that brain pathologies are highly heterogeneous, often requiring tailored solutions depending on the nature of the pathology. For example, diffuse pathologies such as Alzheimer’s disease can be addressed using more specific or age appropriate priors (Fillmore et al., 2015), as it causes global atrophic changes without distorting the general shape of the brain.

In contrast, for focal lesions, the appropriate solution depends on the type of affected tissue and size of the lesion. For example, small white matter lesions typically only have a minor effect on brain mapping but can bias subsequent analyses such as voxel-based morphometry (VBM) (Guo et al., 2019). Such an impact can be mitigated using lesion filling where the lesioned voxels are replaced with intensity values consistent with neighboring unaffected voxels (Battaglini et al., 2012; Chard et al., 2010; Griffanti et al., 2016; Guo et al., 2019; Magon et al., 2014; Popescu et al., 2014; Prados et al., 2014; Schmidt et al., 2019). Larger gray matter lesions (e.g. large focal cortical dysplasia) cannot be addressed using lesion filling unless this is constrained to replicate only healthy gray matter intensities and preserve cortical shape. Pathologies affecting different brain tissues, i.e. not confined to gray matter, white matter or a CSF compartment such as brain tumors, stroke, tumefactive multiple sclerosis (MS) and others are more difficult to tackle. Moreover, the presence of pathological mass effect, and perilesional edema adds to this difficulty. Such pathology can potentially degrade the performance of many steps in a brain mapping pipeline, e.g. skull stripping (Iglesias et al., 2011; Isensee et al., 2019; Lutkenhoff et al., 2014), making it difficult and even impractical to use automated whole-brain structural mapping pipelines for such patients. Freesurfer(Fischl, 2012) recon-all (FreeSurferWiki, 2020a) failure rates can be as high as 30% (Reid et al., 2016) in moderately pathological cerebral palsy brain images, and even higher in case of brain tumors (Zhang et al., 2017).

A number of methods have been proposed to address the problem of brain mapping in the presence of pathology. However, to date there is no consensus on which are the most optimal, nor is a solution available that can be broadly applied to different types of lesions. For example, in case of small or subtle lesions Freesurfer (Fischl, 2012) recon-all (FreeSurferWiki, 2020a) and similar pipelines may still work. Larger or more obvious lesions may benefit from a cost function masking (CFM) approach (Andersen et al., 2010; Brett et al., 2001). This means that the lesion is masked out of the input brain image, and the registration is limited to the non-lesioned brain tissue. CFM has been reported to improve results in some cases (Andersen et al., 2010; Kim et al., 2007), but may also result in a lower quality parcellation in case of large lesions (Nachev et al., 2008). Others resort to functional mapping using resting-state functional MRI to parcellate all healthy gray matter (Wang et al., 2015), or use task-based fMRI for a partial mapping (Reid et al., 2016). Few studies have exclusively investigated this issue in different pathological cohorts (Ledig et al., 2015; Nachev et al., 2008; Solodkin et al., 2010; Weiller et al., 1995). Besides the CFM approach, only two of these resulted in dedicated solutions that were made available non-commercially, namely enantiomorphic normalization (Nachev et al., 2008) http://www.bcblab.com/BCB/Normalisation.html and MALP-EM (Ledig et al., 2015) https://github.com/ledigchr/MALPEM.

## 2. Aims of the study

First, we propose a new workflow to reliably segment/label MR images in the presence of a broad spectrum of focal brain pathologies, whether extra-axial or intra-axial, including large, bilateral, heterogenous lesions with and without mass effect. In doing so, we provide a solution that facilitates state-of-the-art MRI analyses in clinical populations. Subsequently, we test and evaluate its performance in a sample of clinical data and a sample of realistic synthetic data.

## 3. Material and Methods

### 3.1. Proposed workflow

We present a fully automated open-source image processing pipeline called “Virtual brain grafting” (VBG). Briefly, VBG encompasses two approaches for intra-axial lesions both of which use synthetic donor brain images generated for each subject by the workflow. The first approach is intended for unilateral lesions (uVBG) and generates a donor brain using the native non-lesioned hemisphere and one synthetic hemisphere. For bilateral lesions (bVBG), a synthetic brain image is used directly as a donor brain. Both approaches use 2 mm full-width half-maximum (FWHM) 3D Gaussian smoothed masks to fill the lesion.

A third approach, inspired by the recent work of Hou et al (Hou et al., 2020) is included for extra-axial lesions, where the lesion is zero-filled, though this is not based on virtual grafting. VBG uses various open-source Unix-based image processing programs in a fully automated pipeline, including ANTs (Avants et al., 2011) v2.1.0, FSL (Jenkinson et al., 2012) v6.0, MRtrix3 (Tournier et al., 2019) v3.0_RC3 and Freesurfer (Fischl, 2012) v6.0. The steps outlined below may be executed with different programs; however, VBG as described here is fully automated and available for use without the need for any closed-source software. Required inputs are a 3D T1-weighted image and a binary integer format lesion mask covering the entire pathology in the same space, which can be obtained using any manual or semi-automated segmentation tool-box (McCarthy, 2020; Yushkevich et al., 2006).

VBG can be split into two main parts, i.e. (1) lesion filling and (2) whole-brain parcellation of the lesion-free output image using Freesurfer (Fischl, 2012) recon-all (FreeSurferWiki, 2020a). The different steps are explained broadly below, illustrated in **Figure 1** and with additional detail provided in the supplementary material.

**Figure 1:**
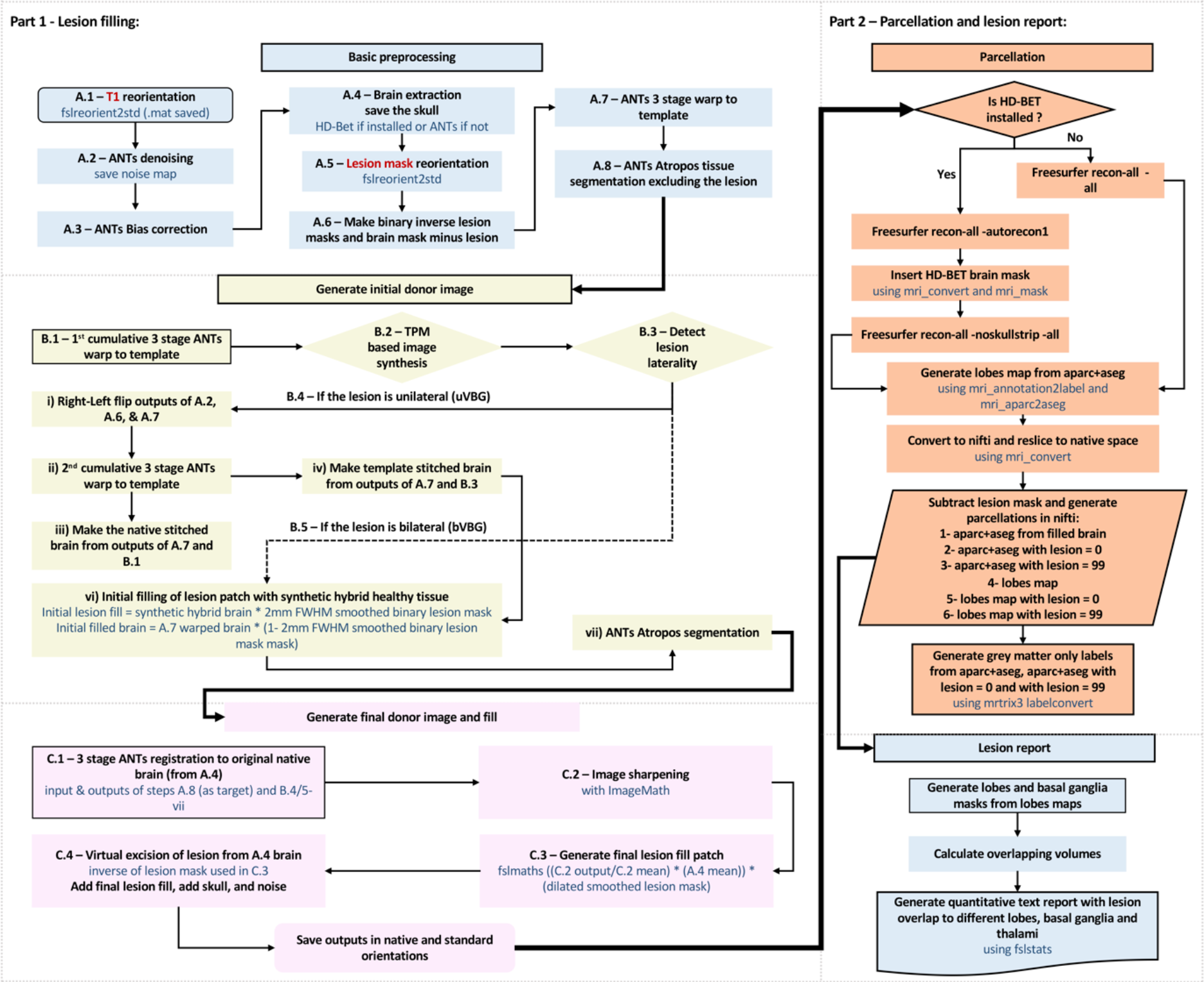
Schematic representation of VBG: Part 1 generates the lesion-free image. It starts in the top left with basic processing and Atropos segmentation (light blue), donor image generation and initial filling (yellow), then final lesion filling (pink). Part 2 is concerned with parcellation (orange). Lastly a text report is generated detailing lesion overlap with various labelled brain structures.

Part 1, lesion filling; this generates the lesion-free T1-weighted images through the following 3 stages:

#### A. Basic preprocessing

This stage applies basic preprocessing to the input T1-weighted image and lesion mask. It starts with image reorientation in FSLpy (McCarthy et al., 2020), denoising (Manjón et al., 2010) and bias correction (Tustison et al., 2010) in ANTs (Avants *et al*., 2011), and brain extraction using HD-BET (Isensee et al., 2019) or ANTs (Avants *et al*., 2011) if the former is not installed. The brain is then warped using cost-function masking in ANTs (Avants *et al*., 2011) to the VBG template brain in MNI space. Finally, ANTs (Avants *et al*., 2011) Atropos (Avants *et al*., 2011) is applied for segmenting the warped brain while excluding the lesion using the brain mask with the lesion subtracted.

#### B. Initial donor image generation

The second stage involves flipping the brain along the right-left axis, iterative deformation to match the template brain using ANTs (Avants *et al*., 2011), synthesizing an initial donor image from the inverse warped template and tissue probability maps (TPMs), and selecting the appropriate pipeline for unilateral or bilateral lesions.

*TPM-based T1-weighted image synthesis (supplementary figure 1)* uses the inverse warped template, prior TPMs, subject’s brain image and TPMs with the lesion excluded. First the input brain images are normalized by a two-pass mean division, and the template (source) TPMs are binarized at a lower threshold of 0.1 and used as masks to isolate each tissue from the source image. The target TPMs are binarized at 95% probability and used to calculate target tissue specific mean intensity with mrstats (Tournier et al., 2019). Each source tissue map is multiplied by the corresponding target tissue mean, and a forced correction of CSF signal intensity is used to scale its maximum to 0.2 of the gray matter mean signal intensity. Finally, all tissues are combined using scalar addition with ImageMath “addto-zero” (Avants et al., 2011).

*Automated lesion laterality detection (supplementary figure 2)* sets the workflow to be followed for the rest of the script. It uses the lesion mask in MNI space, and binary masks of the template’s right and left hemispheres.

If the lesion is unilateral, the lesioned hemisphere is replaced with a synthetic hemisphere retaining the non-lesioned native hemisphere. The resulting image is called a stitched brain. An initial filled brain image is generated by replacing the lesion with healthy tissue from this stitched brain. Next, the stitched brain is further deformed to match the initial filled one, then segmented into different tissues with ANTs (Avants et al., 2011) Atropos (Avants et al., 2011). If the lesion is bilateral the synthetic brain is directly used to derive the initial filled brain, which is then segmented with Atropos (Avants et al., 2011).

#### C. Final donor image generation and fill

This constitutes the last stage in lesion filling. Here the initial filled brain is warped back to native space and sharpened with ANTs (Avants et al., 2011) to create the final donor brain image. The lesion replacement graft is harvested using a 2mm FWHM smoothed mask and inserted into the recipient image (the reoriented input brain). The skull and noise are added back to generate a realistic lesion-free whole head T1 WI. Finally, the initial transformation is reversed to generate the image in native orientation. Whole head, brain extracted, and brain mask images are saved in original and standard orientations.

Part 2, brain parcellation: This involves running recon-all (FreeSurferWiki, 2020a) on the output lesion-free image in native orientation. First recon-all (FreeSurferWiki, 2020a) is run until the brain extraction stage, then the VBG generated brain mask is applied to the recon-all (FreeSurferWiki, 2020a) results, after which it is restarted and run to the end. Finally, the real T1 map and the lesion mask are converted to .mgz, the output aparc+aseg parcellations are converted to .nii, a version is created with a zero-filled lesion patch (aparc+aseg_minL.nii.gz), and another with the lesion mask given the value 99 (aparc+aseg+L.nii.gz), which remains unused in the Freesurfer (Fischl, 2012) v6.0 lookup table. Parcellations are also converted with MRtrix3 (Tournier et al., 2019) and a quantitative text report is generated for the lesion volume and overlap with different lobes, and other structures, e.g. thalami.

### 3.2. VBG testing and evaluation

#### 3.2.1. General outline

Dice similarity coefficient (DSC) calculations represent one of the mainstays for quantitative analyses of segmentation accuracy in the presence of a ground truth. In the case of brain pathology, a manual delineation is typically used as the ground truth. However, for the whole brain this process would be highly time consuming and was not feasible for the current work. Thus, we resorted to two approaches: first, we processed and parcellated T1 WIs from 10 clinical patient participants (real-patients) with gliomas, with and without the proposed method. We included patients with gliomatous lesions of different sizes and locations. A group of healthy control (HC) volunteers (N=10) was also included. We generated a synthetic cohort (N=200), consisting of two groups. First, a lesion-free group (N=100), which was created by non-linear deformation of the HC images to match the mass effect of the patients, referred to henceforth as the synthetic-mass-effect group. Second, a synthetic-patients group (N=100) were generated containing both the mass effect and the lesions. All synthetic-mass-effect images were parcellated with recon-all (FreeSurferWiki, 2020a) and the synthetic-patients images were parcellated after VBG filling. We also attempted to parcellate the real-patients and synthetic-patients’ images without VBG. **Figure 2** shows a schematic of both the image processing steps and the evaluation procedures.

**Figure 2:**
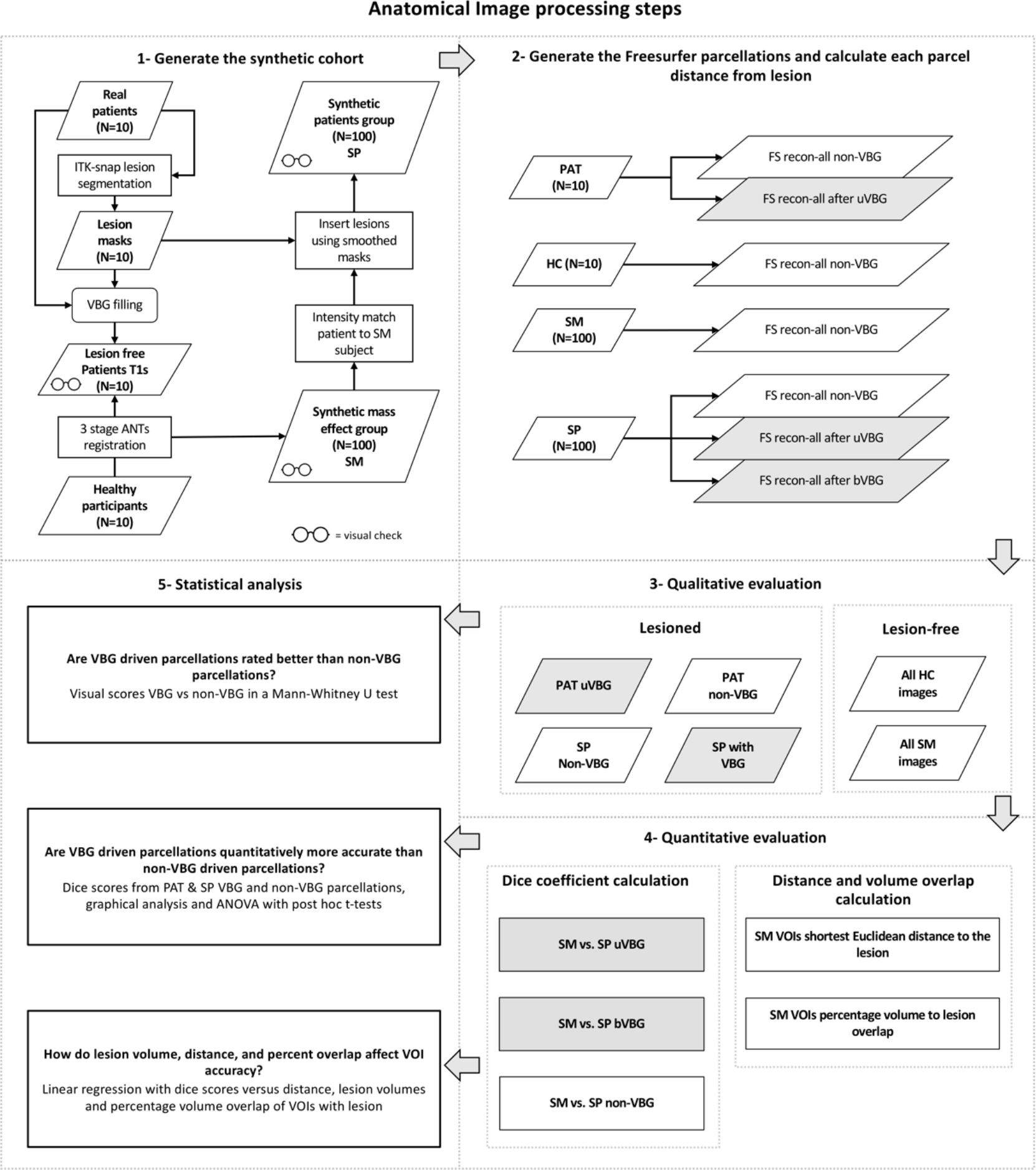
A schematic representation of the VBG validation process. Top: image processing steps applied to generate the Freesurfer parcellations used for evaluation, 1 – 2. Bottom: qualitative and quantitative evaluation and statistical testing, 3 – 5. (PAT – real patients, RH real healthy participants, SM – synthetic-mass-effect group, SP – synthetic-patients group, uVBG – unilateral VBG, bVBG – bilateral VBG), VBG derived datasets are highlighted in gray.

All completed non-VBG parcellations (HC, real-patients, and synthetic-mass-effect), real-patients’ non-VBG uVBG, synthetic-patients non-VBG and corresponding synthetic patients’ uVBG parcellations were qualitatively evaluated (section 3.4.1.). For the synthetic-mass-effect parcellations this was used to confirm acceptable quality prior to further analyses. None of these were excluded. Consequently, all completed synthetic-patients’ parcellations (non-VBG, uVBG and bVBG) could be quantitatively evaluated with DSC using the corresponding synthetic-mass-effect parcellations as the ground truth (section 3.4.2.).

Statistical tests used the results of both evaluation approaches to investigate the accuracy of VBG-driven parcellations. Exploratory analyses using linear regression were also used to investigate the effects of the lesions on each VOI’s DSC in the VBG-driven parcellations (section 3.3.4). **Figure 3** shows representative slices from the patients and uVBG-generated lesion-free output.

**Figure 3:**
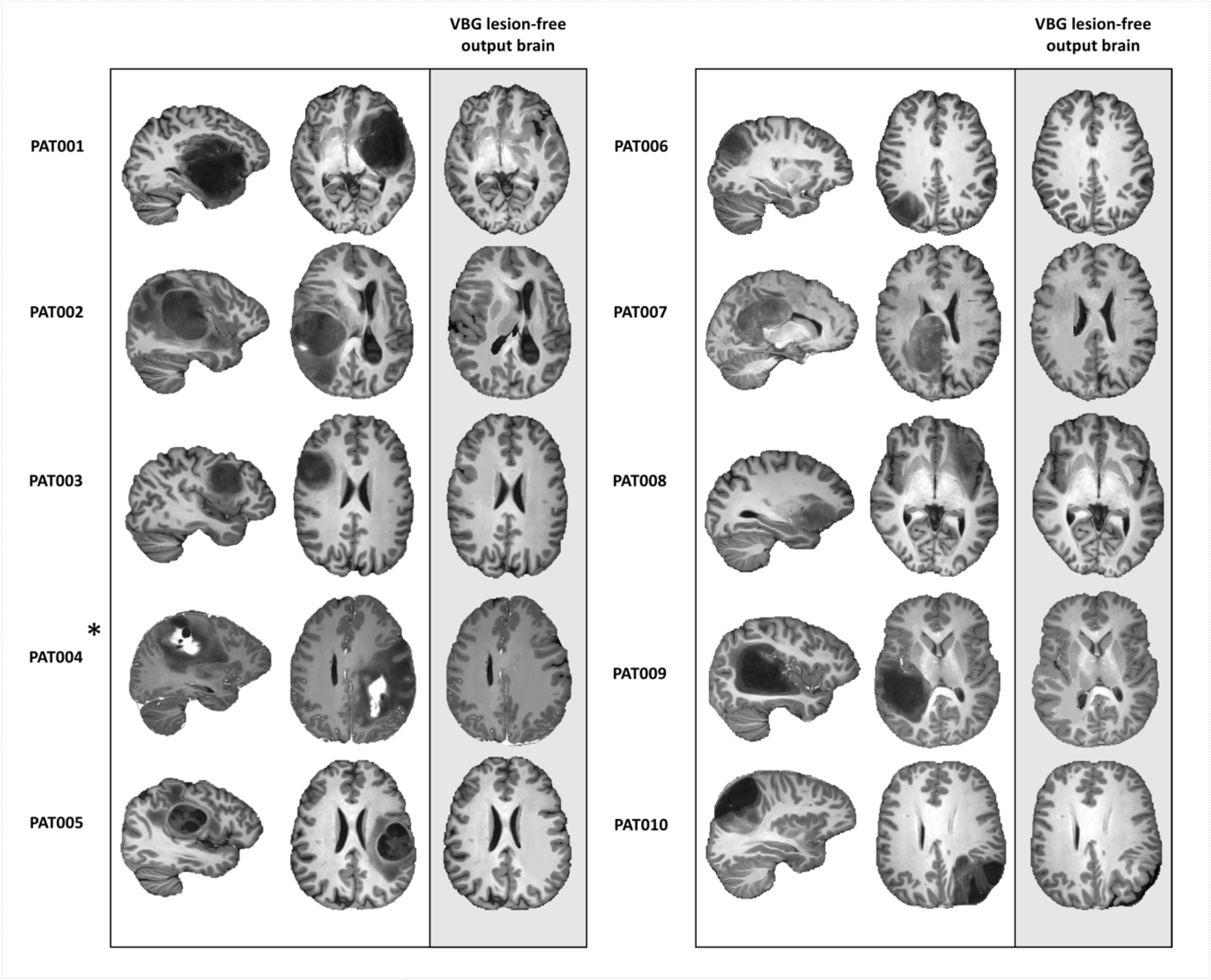
Representative sagittal and axial slices from each patient and the results of uVBG lesion filling demonstrated in axial. Asterisk indicates postcontrast T1 WI used as input.

##### Participants

This study included ten adult surgery naïve patients with gliomas (3 females, 7 males, age range 19 – 61 years, median 36.5), and 10 adult healthy participants (8 females, 2 males, age range 18 – 66 years, median 35) who were free of any neurological disease. This study was approved by the local research ethics committee at UZ/KU Leuven, Leuven, Belgium, study number S61759, and conducted in accordance with the Declaration of Helsinki. A written informed consent was acquired from each participant prior to scanning. **Table 1** lists patient details, MR acquisition, lesion mask volumes and severity of pathological mass effect.

**Table 1.**
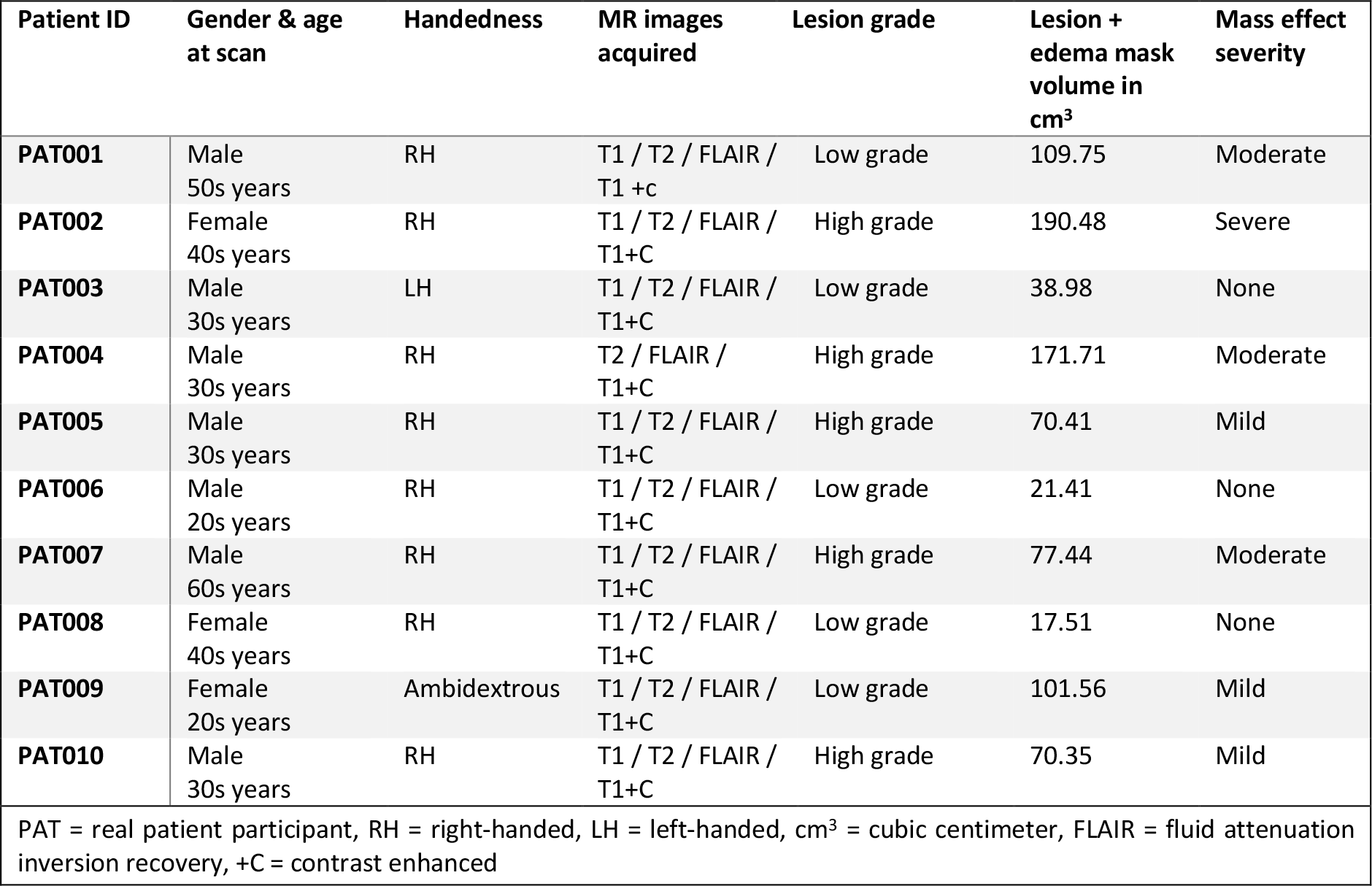
Demographics, handedness, radiological and biopsy results (if available), symptoms at presentation and MR images acquired for each of the patients:

##### Medical image acquisition

MRI acquisition used two 3-Tesla scanners (Achieva dStream and Ingenia-X, Philips Medical Systems, Best, The Netherlands), both with 32 channel phased array receive head coils. The following images were acquired for the patients: sagittal 3D TFE T1-weighted images pre and post intravenous gadolinium injection (TR/TE/FA: 9ms/4.1ms/8°, voxel size: 0.9 × 0.88 × 0.88 mm, matrix: 200 × 288 × 288), sagittal 3D TSE T2-weighted images (TR/TE/FA: 3000ms/280ms/90°, voxel size: 0.9 × 0.98 × 0.98 mm, matrix: 220 × 256 × 256), and sagittal 3D TSE T2 fluid attenuation inversion recovery (FLAIR)-weighted images, (TR/TE/TI/FA: 2800ms/340ms/1650ms/90°, voxel size: 1.0 × 0.98 × 0.98 mm, matrix: 200 × 256 × 256). We used all available modalities for lesion segmentation and only the pre-contrast T1-weighted images for the rest of this study. The healthy participants were scanned on the same scanners with the same non-contrast enhanced T1-weighted scan but reconstructed at 0.6×0.6×0.6 mm voxel size and matrix size of 228 × 384 × 384. One patient (PAT004) was included but had only post-contrast T1-weighted images acquired with the same protocol as the healthy participants.

### 3.3. Image processing

#### 3.3.1. Lesion segmentation

Lesion masks were generated by a neuro-radiologist (AR) in ITK-snap (Yushkevich et al., 2006) v3.8.0 in Mac OSX 10.13.6. We followed the protocol described in (Yushkevich et al., 2019) for multi-modality semi-automated lesion segmentation. However, we aggregated the different lesion tissue components into a single binary mask, which is required for VBG. The remainder of the work used the original unprocessed pre-contrast T1-weighted images. The pathological mass effect of the real patients was subjectively rated (by AR) into none, mild, moderate, and severe for descriptive purposes.

#### 3.3.2. Initial VBG application

Following lesion segmentation, the uVBG approach was applied to generate lesion-free T1 brain images from the real-patients’ data. None were excluded upon visual inspection. All images were included with their original resolution since VBG is designed to accommodate varying spatial resolutions. The lesion-free images generated here were used to create the synthetic cohort.

#### 3.3.3. Synthetic cohort creation

##### The synthetic-mass-effect group

This was generated using ANTs (Avants et al., 2011) nonlinear warping of each HC T1 brain image to each real-patient’s lesion-free uVBG output image, then applying the generated transforms and warps to the whole head T1-weighted images resulting in 100 synthetic subjects. All 10 HCs were registered to each real-patients image mimicking the pathological mass effect of each patient in the 10 HCs. Resulting images were used as the synthetic ground truth for quantitative evaluation. We hypothesized that in the absence of a focal pathology, Freesurfer (Fischl, 2012) recon-all (FreeSurferWiki, 2020a) would be able to run without failure and accurately represent the mimicked pathological mass effect in the parcellations.

##### The synthetic-patients group

These images were generated as follows: first the intensity histogram of each real patient’s original T1 brain image was matched to the target synthetic-mass-effect image with mrhistmatch (Tournier et al., 2019b). The lesion patch and edema were isolated from the patient’s image and inserted into the synthetic-mass-effect brain using a smoothed lesion mask (2 mm FWHM 3D gaussian kernel) to avoid a sharp interface with the recipient image. This expanded our test group to 100 simulated patients, accurately representing each patient’s pathology in each of the 10 healthy volunteers (supplementary figure 4).

#### 3.3.4. Parcellation

All VBG lesion filling and recon-all (FreeSurferWiki, 2020a) parcellations were done on a dedicated compute node of the Vlaams (Flemish) Supercomputer Center (VSC) with two Intel Xeon Gold 6240 2.6 GHz CPUs, 36 cores in CentOS 7.8.2003. The HC (N=10) and synthetic-mass-effect (N=100) data were parcellated using recon-all (FreeSurferWiki, 2020a) with an HD-BET (Isensee et al., 2019) brain mask inserted, as done in VBG. The real-patients (N=10) and synthetic-patients (N=100) data were parcellated with uVBG and attempted after only zero-filling the lesion patch similar to CFM, and also without VBG filling (non-VBG). bVBG was applied to a subset of synthetic-patients (N=25), since there were no true bilateral lesions in our sample. Non-VBG real-patients and synthetic-patients parcellations were used as control parcellations to compare recon-all (FreeSurferWiki, 2020a) success/failure rates and parcellation quality with and without VBG. A total of 455 recon-all (FreeSurferWiki, 2020a) analyses were attempted, each with a runtime cap of 8 hours using GNU timeout. Those that quit with an error or exceeded the timeout duration were considered to have failed, and all parcellations were allowed two attempts in case of failure. We imposed the 8-hour time limit after an initial test of non-VBG recon-all (FreeSurferWiki, 2020a) on the first 18 SP images using 2 cores per subject continued for 24 hours with less than 50% completion rate, while VBG driven recon-all tended to finish in under 7 hours.

### 3.4. Evaluation

#### 3.4.1. Qualitative evaluation

Two independent experienced neuro-radiologists (NS & SS) visually evaluated the parcellations and assigned a quality score. We attempted to minimize rater bias, particularly for the evaluation of lesioned brain parcellations. More specifically, our main concern was blinding the raters to whether the parcellation came from VBG or not, and whether the source was real or synthetic data. To this end we resorted to (a) coding and mixing datasets together, and (b) standardizing the parcellations by using the lesion free T1-weighted images as underlays and subtracting the lesion patch from all parcellations except the HC group, which we used to imply a gold standard. This allowed us to blind our expert raters to the source image of all parcellations of interest. Images were given to the raters as high-resolution multi-frame panels in axial, coronal and sagittal with 10 mm interslice gap, generated using fsleyes render (McCarthy, 2020). We used Gwet’s AC2 (gamma) (Gwet, 2016) as a measure of inter-observer agreement for the parcellations evaluated by both experts using Matlab r2018a and mReliability tools (Girard, 2016). Example coronal slices from each qualitative score are shown in **figure 4**.

**Figure 4:**
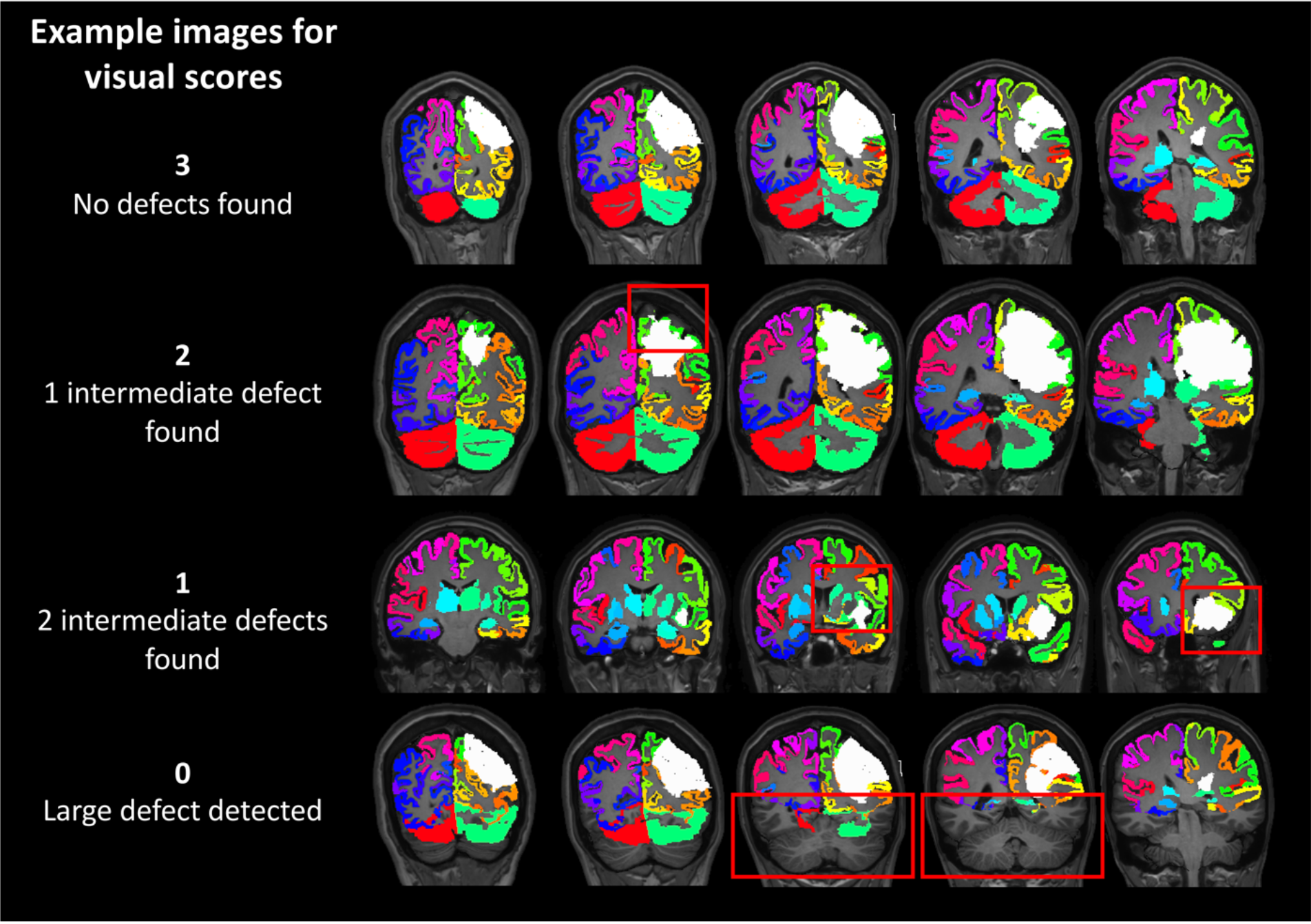
Representative coronal slices for each qualitative score (Red boxes indicate identified defects). Score 3 indicate the best quality and 0 indicates the worst.

The qualitative evaluation protocol we used can be described as follows:

A defect (error) may be minor, intermediate or major. A minor defect was defined as an unlabeled cluster of voxels (e.g. 10) belonging to gray matter, or non-brain tissue e.g. dura labelled as gray matter, or a more subtle focal underestimation of the cortical ribbon thickness. An intermediate defect/error was a larger falsely labelled or unlabeled area on the scale of the inferior frontal gyrus pars orbitalis, or anterior temporal pole. Finally, a major defect was any defect on the level of a whole gyrus or larger. We defined four categories for the quality of a parcellation: 3 – Good, up to 3 minor unconnected defects, which can happen with structurally normal data (FreeSurferWiki, 2020b; Guenette et al., 2018; Klein et al., 2017, 2005; McCarthy et al., 2015). 2 – Acceptable, up to 5 minor, unconnected, or 1 intermediate scale defect/error. 1 – Fair, between 5 and 7 unconnected minor defects, or 2 intermediate unconnected defects. 0 – Poor, > 7 unconnected minor defects, > 2 intermediate defects or a major defect.

Experts rated all completed non-VBG synthetic-patients, real-patients parcellations, corresponding uVBG driven ones, all the synthetic mass-effect, and HC parcellations. We also added a group of a-posteriori warped HC parcellations (HC*), where each HC parcellation was warped to a patient using ANTs (Avants et al., 2011) and nearest-neighbor interpolation.

#### 3.4.2. Quantitative evaluation

We calculated DSC in ANTs (Avants et al., 2011) between each pair of VOIs present in the synthetic-patients and synthetic-mass-effect parcellations (ground truth) for all completed uVBG, bVBG and non-VBG parcellations. For reference, DSC is a measure of spatial similarity between two binary images, ranging between 0 and 1, with 0 meaning no similarity and 1 meaning perfect agreement. DSC can be expressed as follows (if X represents the synthetic-mass-effect parcellation VOI and Y the synthetic-patient’s):

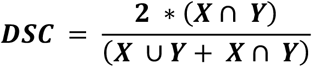

In Matlab r2018a we calculated the center of mass (CoM) of each label in the synthetic-mass-effect parcellations, then the shortest Euclidean distance from each VOI’s CoM to the lesion mask. This is referred to simply as “distance”. Lastly, we calculated the percent of volume by which each VOI overlaps with the lesion mask, referred to for simplicity as “percent overlap”, another factor we hypothesized would influence accuracy.

#### 3.4.3. Statistical analyses

To ensure the validity of the synthetic-mass-effect group parcellations we first compared their visual scores to those of the HCs plus the a-posteriori warped HC parcellations using an unpaired Mann-Whitney U test. Secondly, we asked “Are *VBG driven parcellations qualitatively rated higher than non-VBG parcellations?”* To answer this question, we compared the ratings of the real-patients and synthetic-patients non-VBG driven parcellations to corresponding unilateral VBG driven parcellations using a paired Mann-Whitney U test.

The remaining analyses focused on the synthetic-patients quantitative evaluation results. Here, we asked “*Do VBG driven parcellations have higher dice scores than non-VBG driven parcellations?”* All dice values from the three parcellation types were plotted for visual comparison. Next, we compared the mean dice values from the three parcellation methods. First, we compared completed non-VBG parcellations to the corresponding uVBG (N = 48 non-VBG and N = 48 uVBG) and bVBG (N = 25 non-VBG and N = 25 bVBG) ones separately using paired two-sample t-tests. Then the common parcellations from the three approaches in a one-way ANOVA and post hoc paired t-tests (N = 20)

Finally, we explored the effect of the lesion on VBG DSC with Spearman correlations using the lesion volumes, and average DSC, using the lesion volume acquired from the real-patients image for each 10 synthetic-patients images containing the same lesion. Next, we applied linear regression to investigate the effect of different lesion properties on DSC, namely lesion volume, distance of the VOI from the lesion in mm, and percent overlap between the lesion and the VOI. We hypothesized that larger lesions would result in a global reduction of DSC scores, and that VOIs closer to the lesion and/or with higher lesion overlap would have a lower DSC score than those further away and/or without overlap. The parameters explaining most of the variance were also used in a stepwise multiple linear regression model, with the pooled VOI DSC scores from all subjects in the synthetic-patients group as dependent variable. These tests were applied to uVBG and bVBG DSC values separately in Matlab r2018a.

## 4. Results

First, the parcellations derived from the synthetic-mass-effect group were comparable in visual quality scores (88 scored 3, 11 scored 2, and 1 scored 1) to the HC group and a-posteriori warped HC parcellations (15 scored 3, and 5 scored 2). A two-tailed Mann-Whitney U test comparing both groups without pairing indicated no significant differences, U(100,20) = 1127.5, z = 1.486, *P* = 0.137. Secondly, Gwet’s AC2 (Gwet, 2016) was used to measure inter-observer reliability. This showed a good overall agreement between both experts using all common parcellations (gamma = 0.943, observed agreement = 0.960, chance agreement = 0.318). Thirdly, we report on the results of parcellation, qualitative and quantitative evaluations, as well as the paired statistical comparison of parcellation methods and exploratory analyses (see supplementary information for results of the unpaired comparison).

We included the results from the second attempt (non-VBG, N=48/52 completed/failed, grand average DSC = 0.675, and standard deviation = 0.281) due to higher DSC values and more completed parcellations (see supplementary information for results of the first attempt). All synthetic-patients uVBG parcellations succeeded on the first attempt. Only 1/10 Patient’s uVBG parcellation failed initially, but completed on the second attempt, compared to 6/10 failed after zero-filling and 4/10 failed non-VBG patients parcellations. We used the minimum of the visual scores assigned for the common parcellations by both experts, and the remaining unique scores from each expert.

*Qualitative evaluation* showed real-patients uVBG parcellations scored significantly higher than corresponding non-VBG ones, U(6,6) = 33, z = 2.738, *P* < .01. uVBG also outperformed non-VBG parcellations in the synthetic-patients group. This was confirmed on a paired U-test comparing corresponding parcellations from both methods U(48,48) = 2076, z = 7.336, *P* < .001. **Figure 5** shows plotted visual scores from all synthetic-patients non-VBG and uVBG parcellations plus the average DSC scores from the synthetic-patients common to the three parcellation approaches. Unpaired analysis results are detailed in supplementary information.

**Figure 5:**
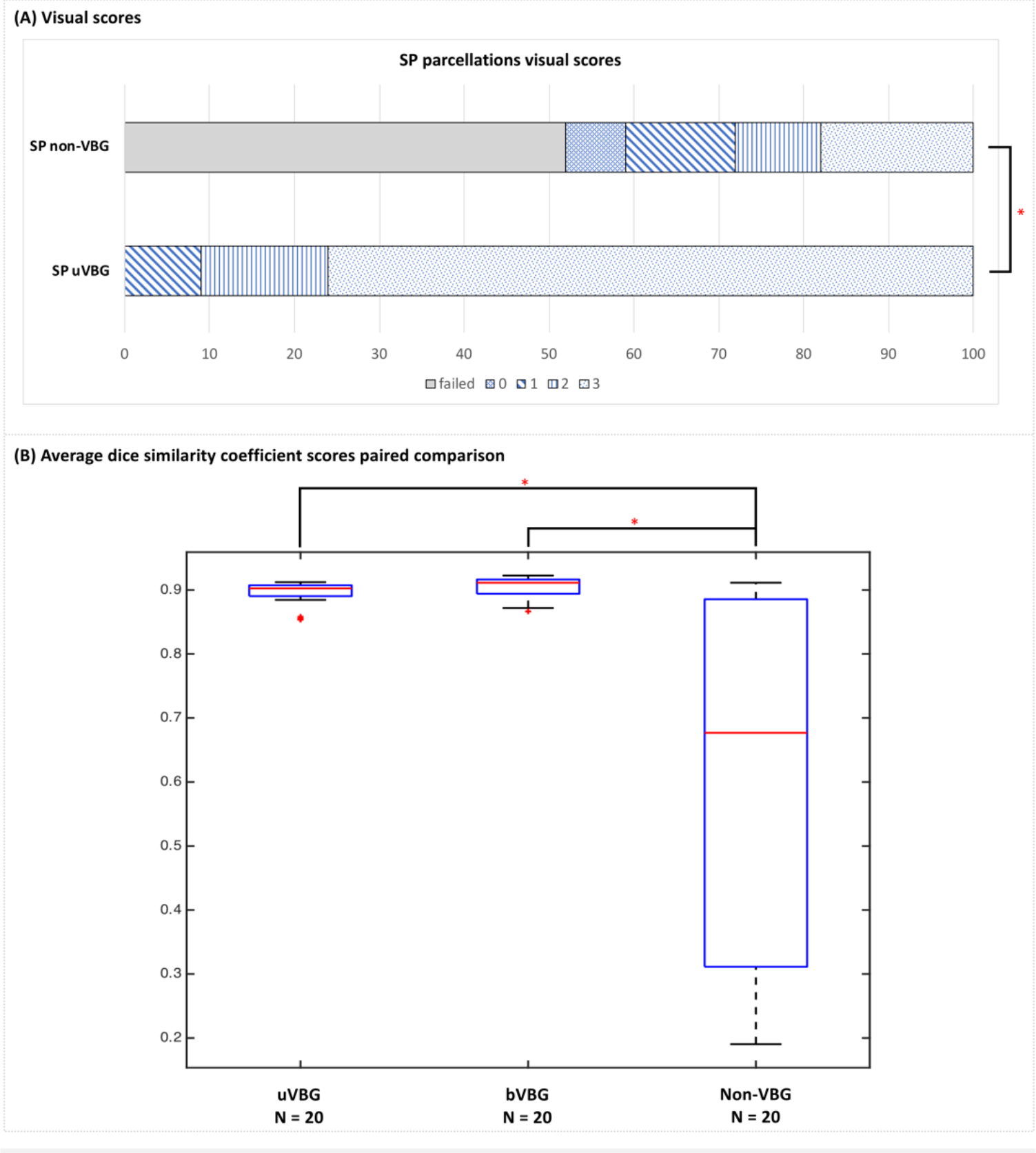
(A) shows synthetic-patients (SP) visual scores for uVBG & non-VBG parcellations. (B) shows box plots of grand average dice scores per parcellation approach, for paired (only commonly parcellated by the 3 approaches) comparison, asterisks indicate statistical significance at *P* <.01, error bars indicate standard error.

*Quantitative analysis* also confirmed this impression using the synthetic-patients average DSC scores, uVBG out-scored non-VBG in the paired two-sample t-test (N = 48 each, t-stat = 3.011, *P* < .01, t-critical two-tailed = 2.684). One-way ANOVA and post hoc paired t-tests comparing the common parcellations from the three approaches (uVBG, bVBG and non-VBG, N = 20 each) showed significant differences between the three groups (*P* < .001), with both VBG approaches scoring significantly higher than non-VBG.

We found a strong logarithmic relationship between distance and DSC, thus log_10_ scaled distance measure was used for exploratory analyses. Results from the statistical tests used to explore the lesion’s influence on dice scores are listed in **table 4**.

**Table 2.** Attempted, completed and failed parcellations, inclusions for visual QC, and DSC calculations, as well as Gwet’s AC2 results for inter-rater reliability:

| Parcellations | Lesioned data (PAT + SP) |  |  | Lesion-free data (HC + SM) |
| --- | --- | --- | --- | --- |
|  | Non-VBG | uVBG | bVBG | Non-VBG |
| Attempted | 10 PAT + 100 SP | 10 PAT + 100 SP | 25 SP | 10 HC + 100 SM |
| Completed | 6 PAT + 48 SP | 10 PAT + 100 SP | 25 SP | all |
| Failed 1<sup>st</sup> attempt | 6 PAT + 56 SP (zF) | 1 PAT* | none | none |
| Failed 2<sup>nd</sup> attempt | 4 PAT + 52 SP (non-VBG) | none | none | none |
| Visual QC per expert | 6 PAT (common) + 34 SP (24 unique) | 10 PAT (common) + 34 SP corresponding to non-VBG SP (24 unique and 10 common) | none | 10 HC (common) + 10 HC*(common) + 61 SM (39 unique + 22 common) |
| DSC | 48 SP non-VBG vs. corresponding SM | 100 SP vs. corresponding SM | 25 SP vs. corresponding SM | 100 SM (as ground truth) |
| Gwet's AC2 (Γ) scores | PAT non-VBG = 0.910 | PAT uVBG = 0.932 - SP<br>uVBG = 0.849 | - | All = 0.967 - SM only =<br>0.969 - HC only =<br>0.988 - HC* only =<br>0.936 |
| PAT = real patient participant, SP = synthetic patient, HC = healthy control participant, SM = synthetic mass effect subject, uVBG = unilateral VBG, bVBG = bilateral VBG, QC = quality check, DSC = dice similarity coefficient, zF = zero filled, non-VBG = without VBG filling, PAT* = PAT004 (contrast enhanced t1 image), HC* = a-posteriori warped HC parcellations |  |  |  |  |

**Table 3.** (A) Visual scores summary statistics, and (B) paired DSC comparison results

| (A) Visual scores summary statistics |  |  |  |  |  |  |  |
| --- | --- | --- | --- | --- | --- | --- | --- |
| Parcellation | Count | Completed/failed | Visual QC | Score 0 (worst) | Score 1 | Score 2 | Score 3 (best) |
| uVBG PAT | 10 | 10/0 | 10 | 0 | 1 | 1 | 8 |
| non-VBG PAT | 10 | 6/4 | 6 | 0 | 1 | 4 | 1 |
| uVBG SP | 100 | 100/0 | 48 | 0 | 9 | 15 | 76 |
| bVBG SP | 25 | 25/0 | none | - | - | - | - |
| Non-VBG SP | 100 | 48/52 | 48 | 18 | 10 | 15 | 7 |
| Non-VBG SM | 100 | 100/0 | 100 | 0 | 1 | 12 | 87 |
| Non-VBG HCs | 10 | 10/0 | 10 | 0 | 0 | 1 | 9 |
| Non-VBG HCs* | 10 | 10/0 | 10 | 0 | 0 | 4 | 6 |
| (B) Paired DSC comparison results (matching the three approaches) |  |  |  |  |  |  |  |
| Summary statistics |  |  |  |  |  |  |  |
| Groups | Count |  | Average |  | Std dev |  |  |
| uVBG | 20 |  | 0.894 |  | 0.020 |  |  |
| bVBG | 20 |  | 0.903 |  | 0.017 |  |  |
| Non-VBG | 20 |  | 0.617 |  | 0.286 |  |  |
| ANOVA: Single factor |  |  |  |  |  |  |  |
| Source of variation | df |  | F |  | P-value |  | F critical |
| Between groups | 2 |  | 19.188 |  | < .0001 |  | 3.158 |
| Within groups | 57 |  |  |  |  |  |  |
| Total | 59 |  |  |  |  |  |  |
| Post hoc t-tests: Paired two-sample assuming unequal variances |  |  |  |  |  |  |  |
|  | df |  | t stat |  | P (two-tail) |  | t critical (two-tail) |
| uVBG vs non-VBG | 19 |  | 2.871 |  | < .01 |  | 2.093 |
| bVBG vs non-VBG | 19 |  | 3.008 |  | < .01 |  | 2.093 |
| SP = synthetic patient, PAT = patient participant, SM = synthetic mass effect group, HCs = healthy control participants, HCs* = a-posterior warped HCs, uVBG = unilateral VBG, bVBG = bilateral VBG, DSC = dice similarity coefficient, uVBG* = uVBG corresponding to completed non-VBG parcellations, ANOVA = analysis of variance, df = degrees of freedom, Std dev = standard deviation, Stat = statistic |  |  |  |  |  |  |  |

**Table 4.**
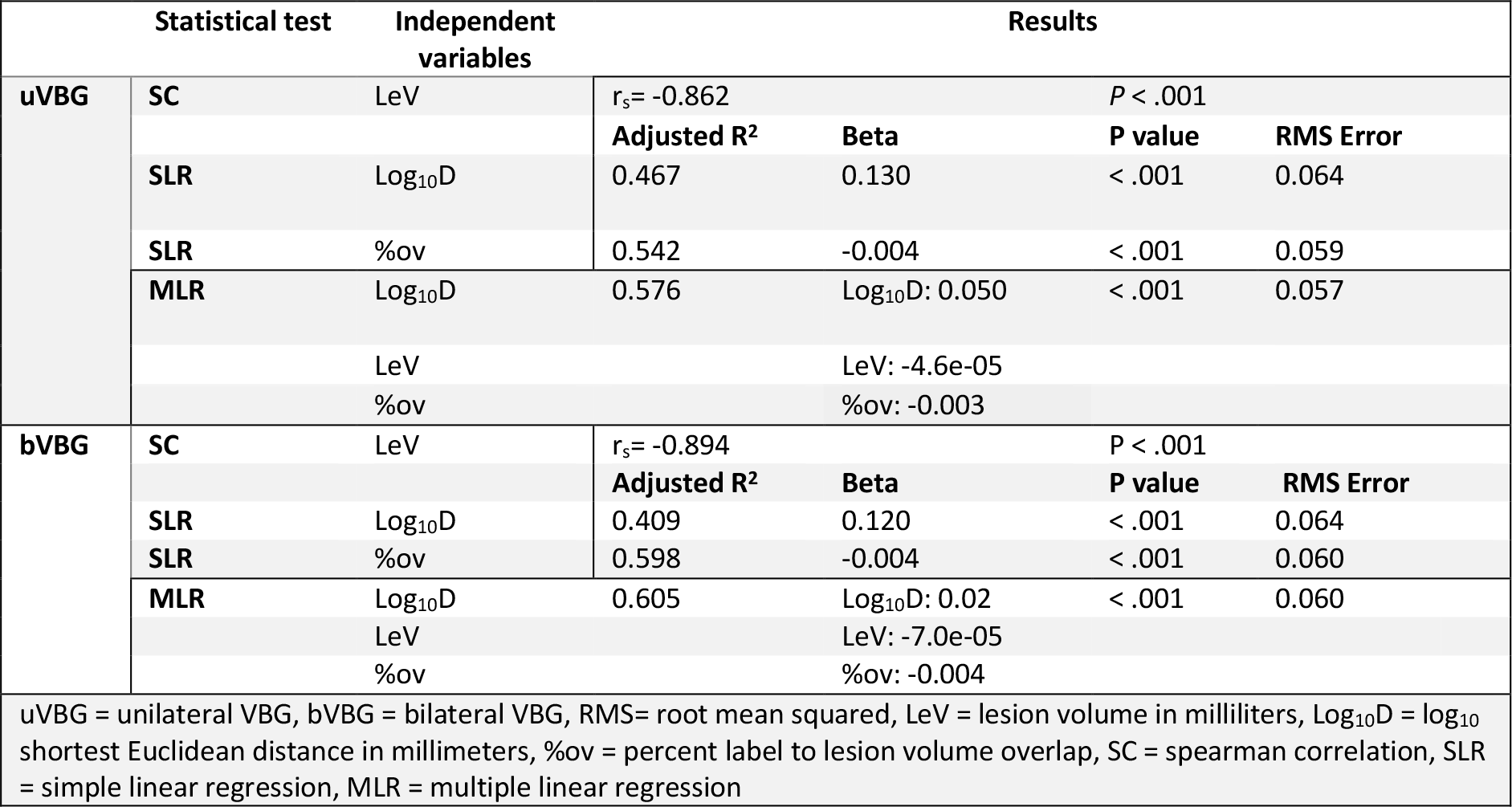
Results of exploratory analyses for uVBG and bVBG using all VOI DSC

Briefly, we found a strong significant inverse association using Spearman correlation (uVBG (N=100): r_s_ = −0.863, *P* < .001; bVBG (N=25): r_s_ = −0.89, *P* < .001), between the average dice scores per subject and lesion volumes. Simple linear regression (SLR) revealed a significant inverse relation between the DSC per VOI and log_10_ scale distance, and a significant inverse relation to percent overlap. Results of the SLR tests for uVBG and bVBG dice scores versus log_10_ distance are shown in **figure 6**. In a multiple linear regression model, these variables together explained 58% of the variance in dice scores.

**Figure 6:**
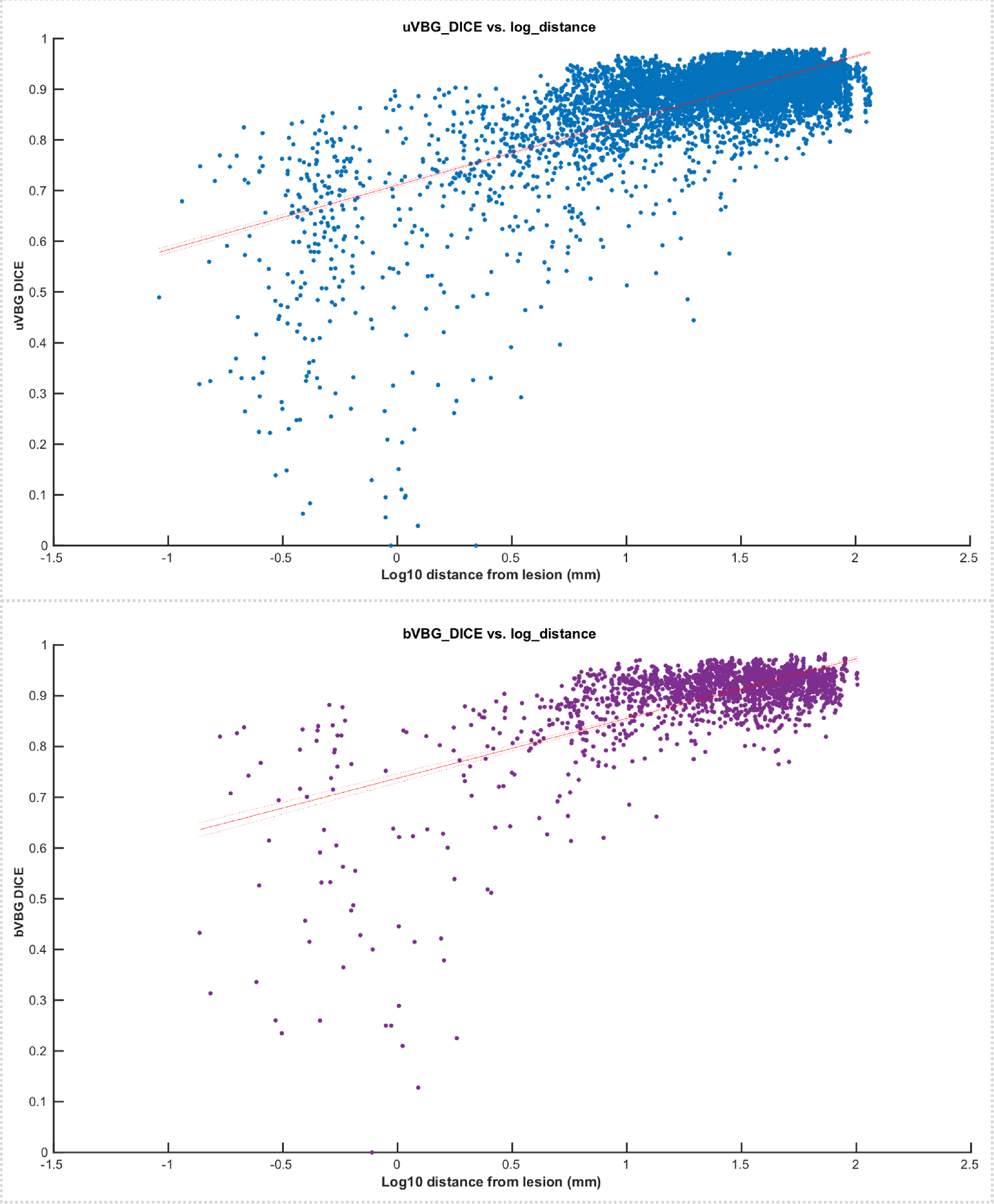
Results of simple linear regression for uVBG (top), and bVBG (bottom) VBG approaches versus log_10_ distance. Red line and parallel dotted range represent the intercept and 95% confidence interval

## 5. Discussion

The first aim of this work was to propose and explain VBG as a workflow for heterogeneous brain lesion filling and optional subsequent structural mapping using Freesurfer (Fischl, 2012) reconall (FreeSurferWiki, 2020a). Our second aim for this study was to test and evaluate the quality and accuracy of the VBG driven whole brain parcellations. We chose a test sample of preoperative patients with gliomas for this work as they provide a variety of lesion sizes, locations, and mass effect.

Our evaluation shows a significant benefit from using VBG in this group, both qualitatively and quantitively in the real and synthetic cohorts. Put simply, VBG allows an accurate parcellation for patients where recon-all (FreeSurferWiki, 2020a) would otherwise fail to complete. Our exploratory analysis partially explained the variation in VOI dice values in the VBG driven parcellations of the synthetic cohort. In intuitive terms, our analysis showed that VOIs lost 0.004 DSC for every 100 mL increase in lesion, .05 DSC was gained for a 10-fold increase in distance to the lesion, and 0.003 DSC is lost for every 1% label volume lost to overlap with the lesion. However, there is an inherent multicollinearity between the three parameters as in the case of larger lesions there is less probability for VOIs to be more distant, and more chance for overlap with the lesion mask.

Only 1 of 135 VBG recon-all (FreeSurferWiki, 2020a) runs failed, a real patient’s postcontrast T1 weighted image that was appropriately filled by VBG but lagged in the automated topographical error correction (FreeSurferWiki, 2020c) of recon-all (FreeSurferWiki, 2020a). We hypothesized that the cause was the gadolinium signal confounding cortical surface morphology. This was confirmed upon inspecting recon-all (FreeSurferWiki, 2020a) inflated surfaces, which showed major defects in the vicinity of the bright blood vessels (supplementary figure 5). Thus, we repeated recon-all (FreeSurferWiki, 2020a) without a time-limit. This finished without error after 18 hours and the dataset was included. Such cases would be better addressed using another modality to assist with the cortical surface reconstruction, e.g. T2 or FLAIR. Alternatively, a different surface reconstruction approach, e.g. FastSurfer (Henschel et al., 2020) could be used, which may probably be adopted in the next version of VBG as this also promises to significantly shorten runtime.

Adequate lesion filling must conform to the healthy brain tissue morphology and image intensity pattern. Furthermore, sharp interfaces between the lesion fill graft and surrounding recipient brain image must be avoided. VBG achieves this by generating a synthetic donor image that matches the input brain image to ensure the filling tissue does not carry concomitant pathologies or parts of the main lesion from the relatively non-lesioned side. In bilateral lesions, there is no healthy hemisphere to rely on, and the unilateral approach would simply duplicate the whole lesion into the filling patch (supplementary figure 6). Thus, the synthetic images are used directly for the initial and final lesion filling. Here, we did not observe a significant difference in performance between unilateral and bilateral approaches, as quantified with mean DSC.

VBG can be used for the parcellation of a wide variety of lesions, both unilateral and bilateral, as opposed to previously described lesion filling based approaches, which were more suited for smaller unilateral lesions. For example, enantiomorphic normalization (Nachev et al., 2008), which relies on the native healthy hemisphere only for patching the lesion prior to normalizing to the MNI152 template brain. It is available non-commercially (http://www.bcblab.com/BCB/Normalisation.html) but only works for unilateral lesions with no significant midline shift (Solodkin et al., 2010) and does not apply a whole brain parcellation afterwards. Another approach called virtual brain transplantation was described in 2010 (Solodkin et al., 2010) also relies on the native healthy hemisphere for lesion filling. These authors also evaluated the accuracy of recon-all (FreeSurferWiki, 2020a) parcellations on the lesion-free images and reported encouraging results in a sample of patients with relatively large stroke lesions. However, to our knowledge this method has not yet been made available on any open-source platforms.

MALP-EM (Ledig et al., 2015) has shown encouraging results in a sample of patients with traumatic brain injury. In contrast to the previously mentioned methods and to VBG, MALP-EM does not need a lesion mask, and does not rely on lesion filling prior to segmentation. It uses multi-atlas matching and expectation maximization to achieve a volumetric segmentation. Thus, theoretically transcending the issue of lesion location and laterality. However, in practice it can still have some errors with larger lesions. Furthermore, MALP-EM uses a different atlas for volumetric mapping, without generating a cortical surface. Structural brain mapping using Freesurfer (Fischl, 2012) enables the standardization of analyses in pathological data to match state-of-the-art methods used in normal data. VBG also enables the use of Freesurfer (Fischl, 2012)-based analyses in such data, e.g. CIFTIFY (Dickie et al., 2019) and the HCP processing pipeline (Glasser et al., 2013). Alternatively, the lesion-free output of VBG could also be used in other structural mapping pipelines such as MALP-EM (Ledig et al., 2015), or ANTs MALF (Wang et al., 2011).

VBG can benefit from a reduction in runtime, and optimizing workflow details such as image synthesis, which can result in occasional expansion of structures within the filling patch in very large lesions (>150 ml), e.g. insula, putamen (supplementary figure 7). The gray - white matter interface may also appear sharper than normal. We tested the bilateral approach (bVBG) in a subset of synthetic data only, as none of the included patients had truly bilateral lesions, only midline shift due to mass effect, or slight contralateral extension through white matter structures (e.g. PAT004). Further testing will investigate VBG’s performance in other pathological populations, e.g. stroke, cerebral palsy, cortical dysplasia, bilateral lesions and in larger samples. Further development will adopt full BIDS (Gorgolewski et al., 2016) compliance, release a VBG docker (Merkel, 2014) image, fine-tune donor image synthesis, reduce run-time, and potentially include different MR modalities, e.g. T2 and FLAIR, from which a T1 image could be synthesized.

Future studies addressing the issue of structural mapping of brain images with large lesions could rely on deep learning solutions, for example a convolutional neural network (CNN) e.g. DeepMedic (Kamnitsas et al., 2017) may be used to segment the lesion, a generative adversarial network can generate a lesion-free version of the input brain, and the last CNN would segment the remaining healthy tissue, e.g. FastSurfer (Henschel et al., 2020), or DeepNat (Wachinger et al., 2017). In this context VBG could benefit deep learning solutions by providing the required ground truth and training data, after sufficient quality checking. This would ideally be undertaken in a multidisciplinary collaborative setup and use different MR modalities from different scanners for maximum yield.

## 6. Conclusions

In this work we have proposed VBG, a new workflow for reliable structural mapping of T1 images using Freesurfer (Fischl, 2012) in the presence of heterogeneous, and large pathologies. Our testing and evaluation show it to be a reliable approach in this group of patients with heterogeneous gliomas, and similarly in the synthetic testing dataset. To conclude, VBG achieves a realistic lesion filling and enables an accurate whole brain parcellation using Freesurfer (Fischl, 2012). It is an open-source workflow, currently requiring no GPU processing, and is available under a Mozilla 2.0 license, via (https://github.com/KUL-Radneuron/KUL_VBG).

## Data Availability

Imaging data in this study are not available for sharing due to patient privacy reasons. The parcellation and statistical data that supports the findings of this study are available for sharing upon request from the authors.
The shell script used for the workflow described is openly available via https://github.com/KUL-Radneuron/KUL_VBG under a Mozilla 2.0 license.

## 7. Acknowledgements

The computational resources and services used in this work were provided by the VSC (Flemish Supercomputer Center), funded by the FWO and the Flemish Government – department EWI. The authors also thank Prof. Josien Pluim, Prof. Steven De Vleeschower, Dr. Laura Michiels for their support, as well as the healthy control participants and participating patients for their participation. LE is supported by the research foundation Flanders (FWO, grant no. G0C0319N) and KU Leuven Sequoia Fund. JB is an aspirant researcher for the FWO (grant no. 11B9919N).

