## Supplementary information for "Virtual brain grafting: Enabling whole brain parcellation in the presence of large lesions"

### Virtual brain grafting supplementary information

The following document includes additional detail regarding the VBG processing pipeline including additional commands to improve its functionality. Several supplementary figures are provided which give further information about the generation of tissue probability maps (SFig1), lesion laterality detection (SFig2), unpaired DSC comparisons (SFig3), synthetic data generation (SFig4) and representative errors (SFig5-7).

#### 1. Supplementary methods

##### Detailed explanation of VBG

The following steps relate to the VBG processing pipeline as outlined in Figure 1 in the main paper.

*A. Basic preprocessing:*

A.1- Image reorientation to standard using fslreorient2std, to align the input T1 WI to the template used by VBG. The transformation matrix is saved and used later on to return to the input orientation.

A.2- Image denoising using ANTs DenoiseImage.sh and save the noise map

A.3- Signal intensity bias correction using antsN4Biascorrect.sh

A.4- Brain extraction using HD-BET[1] or antsBrainExtraction.sh if the former is not installed

A.5- Image reorientation of the lesion mask

A.6- Generate binary inverse (1 - Intensity) of lesion mask, brain mask excluding the lesion and smoothed and dilated lesion masks (we relied on fslmaths with a combination of 2 times mean dilation, smoothing by sigma=2 mm, thresholded at 0.2)

A.7- Three stage (rigid, affine, and SyN[2]) warping to the VBG template brain using antsRegistrationSyN.sh then applying the warps to the mask images and noise maps. This step also generates the inverse warp and inverse warped template image.

A.8- Tissue (GM, WM, & CSF) segmentation of the warped T1 WIs from step A.7 using antsAtroposN4.sh, excluding the lesion patch

*B. Initial donor image generation:*

B.1- First 3 stage antsRegistrationSyN.sh warping to the template brain, generating twice warped images

B.2- TPMs based T1 image synthesis (supplementary figure 1) using the subject specific TPMs from step A.8 and the inverse warped template prior TPMs. The latter set of TPMs are used to fill the defect due to lesion exclusion in the former. Both the target and source images are twice normalized by dividing each by its mean intensity. The filled TPMs are thresholded and binarized and voxel wise populated with tissue specific intensities, matching those of the output of step A.7. This is done by sampling the mean of tissue specific intensities from the target image using a conservative tissue mask (e.g. cortex GM TPM thresholded at top 5% using mrthreshold). The target tissue mean signal value is used to scale up the filled tissue maps to the normalized target intensity, by scalar multiplication, then scaled back up to original intensity values. The resulting tissue intensity maps are then added together (with ImageMath) to generate the first whole brain synthetic image, and any empty voxel are also filled (with ImageMath). Finally, linear histogram matching (with mrhistmatch) to the output of B.1 excluding the lesion is applied and the output is referred to as "1$^{st}$ synth. brain".

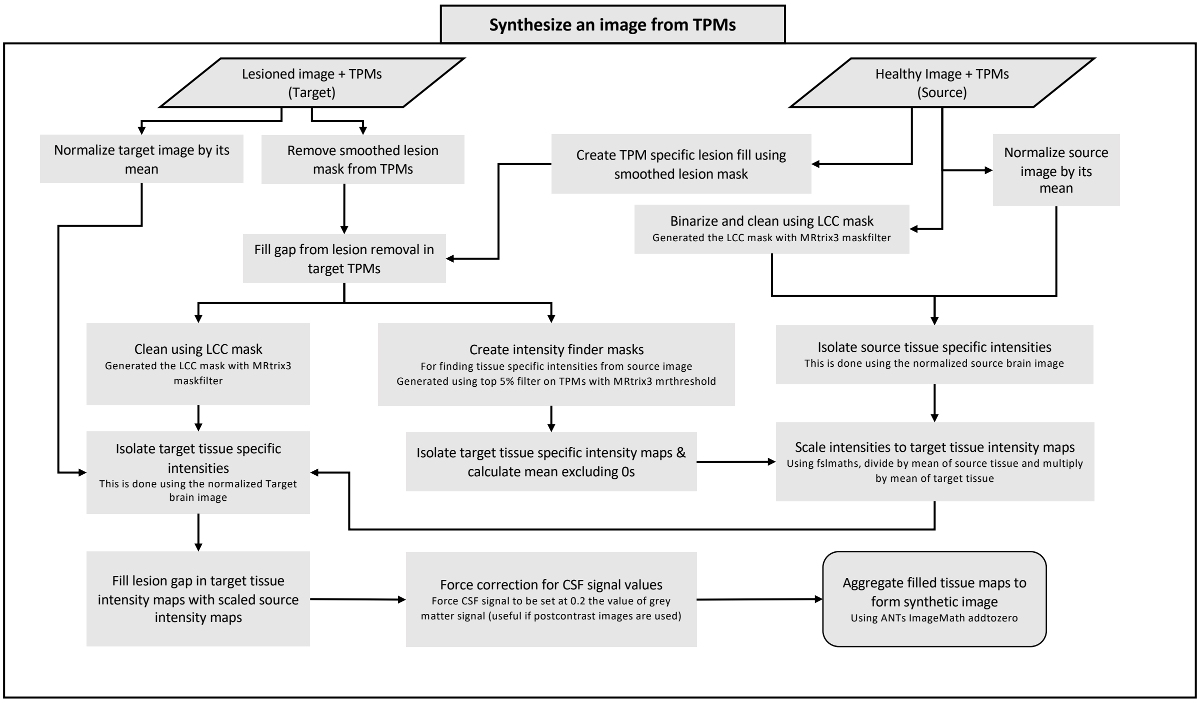


**Supplementary Figure 1:** Schematic showing TPM based image synthesis in VBG

B.3- Automated detection of lesion laterality (supplementary figure 2) sets the workflow to be followed for the rest of the script. It uses the lesion mask in MNI space, and binary masks of the template's right and left hemispheres. First the hemisphere masks are inverse warped to match the subject's brain, then overlap maps between the lesion and each warped hemisphere mask are generated. We calculate a ratio of each hemisphere's lesion burden (rHLb) to the total lesion volume. This can be expressed as follows:

$$rHLb = \frac{Volume(Hemi \cap lesion\ mask)}{total\ lesion\ volume}$$

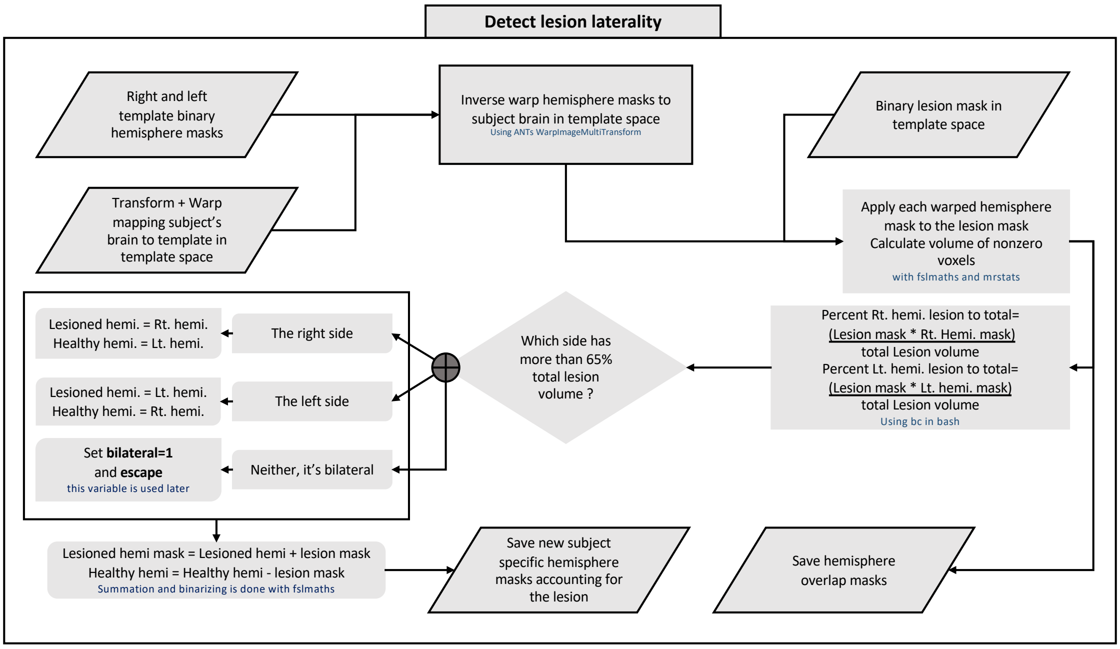


**Supplementary figure 2:** Schematic showing automated lesion laterality detection in VBG

If this ratio is above 65% for one hemisphere, it is identified as the lesioned hemisphere and the unilateral pipeline (uVBG) is followed, if not then the bilateral workflow (bVBG) is applied. Both workflows apply TPM based image synthesis. If the lesion is unilateral, the lesioned hemisphere is replaced with a synthetic hemisphere, retaining the non-lesioned native hemisphere. The resulting image is called a stitched brain. An initial filled brain image is generated by replacing the lesion with healthy tissue from this stitched brain. Next, the stitched brain is further deformed to match the initial filled one, then segmented into different tissues with ANTs [18] Atropos [54]. If *the lesion is bilateral*, no stitched brain image can be generated and the synthetic brain is used instead to derive the initial filled brain, which is then segmented with Atropos [54].

B.4- If the lesion is unilateral:

i. Right-left flipping of the warped brain images, noise maps, and mask images, the outputs of step A.7

ii. Second cumulative 3 stage antsRegistrationSyN.sh warping to the template brain, generating twice warped flipped images and flipped noise map

iii. Warp template hemispheric masks to subject brain in MNI using the transform and warps generated in step B.1. The lesion mask is added to the lesioned hemisphere mask, and subtracted from the healthy hemisphere mask. The subject's brain is then split into a healthy hemisphere, excluding any lesion voxels crossing the midline, and a lesioned hemisphere including any lesion voxels crossing the midline. The latter is discarded and replaced with an image graft from the flipped brain. This creates a synthetic healthy-looking brain image using only the subject's brain, which we refer to as the "stitched native brain". A similar process is applied to the flipped noise map, generating a "stitched noise map".

iv. We repeat the above step using the synth. brain 1 output by step B.3 as the donor instead of the flipped brain, generating a "stitched synthetic brain", which contains a native healthy hemisphere and a synthetic hemisphere.

v. An initial filling of the lesion patch is done using the stitched synthetic brain as a donor, to the output of step B.3 giving an "initial template filled subject's brain".

vi. The output of step B.4-v is then nonlinearly warped (using SyN only) to the output of step B.4-vi. This generates a "stitched to filled lesion free brain".

vii. Tissue segmentation of the stitched to filled lesion free brain using antsAtroposN4.sh.

B.5- If the lesion is bilateral, only step 4-vi is done, using the 1st synth brain image to fill the lesion patch only. The 1st synthetic brain is used instead of the hybrid stitched brain, and the initial filled brain is used instead of the stitched native brain. Then step 4-vii is applied, and step-viii is applied to the initial template filled subject's brain.

*C. Final donor image generation and fill:*

C.1- 3 stage antsRegistrationSyN.sh registration to the native brain (output of A.4). This step uses a brain mask that excludes the lesion from the fixed brain image (CFM).

C.2- Sharpening of the synthetic image with ImageMath SharpenImage.

C.3- The final lesion fill patch is generated with fslmaths, as follows: The C.2 output is divided by its mean then multiplied by the mean of the native brain (from A.4) excluding the lesion, then multiplied by the smoothed lesion mask from A.6.

C.4- The native A.4 brain is multiplied by the inverse of the lesion mask used above, then the lesion patch is added in (using fslmaths). Then, the skull is added back to the filled brain image and finally the noise map is added back. This generates a brain extracted filled image, its brain mask, the denoised filled whole head image and another with the noise added back, which is used as input to Freesurfer. These outputs are generated in original native orientation, i.e. as the image were input, and in standard orientation.

*Part 2, brain parcellation*: This involves running recon-all on the output of step C.4. Recon-all is initiated with the -autorecon1, which runs the pipeline up to the brain extraction step and quits. At this point we insert the brain mask generated by VBG step A.4 by simultaneously converting to ".mgz" format and reslicing it to match the brainmask.mgz using mri_convert. This mask is then applied to the recon-all generated brainmask.mgz file and used to replace the brainmask.auto.mgz file. Next recon-all is restarted, this time omitting the -i flag and using the -all and -noskullstrip. This produces the default recon-all parcellation images, which are converted to nifti using mri_convert. The lesion mask is then removed, generating an aparc+aseg with the lesion voxels set to 0 (aparc+aseg_minL.nii.gz), and another with the lesion patch added back with the value 99 (aparc+aseg+L.nii.gz). These are then converted to contiguous grey matter only parcellation maps using MRtrix3 labelconvert. Lobes parcellation maps are also generated using mri_annotatio2label and mri_aparc2aseg. Finally, a quantitative text report is generated for the lesion volume, lesion to lobe and to basal ganglia overlap statistics.

**Additional options for customization and improved functionality**

VBG was initially developed for unilateral intra-axial neoplastic lesions but has since been extended for and preliminarily applied to other pathologies, as well as bilateral, and multifocal lesions. We also provide a workflow to generate a study specific donor brain from pathological data by combining healthy hemispheres from different subjects to be used as a template/donor image for VBG, called KUL_cook_template_4VBG.sh. VBG also provides the following optional input flags, among which is a pediatric friendly mode. Optional arguments to the VBG script are listed below:

- "-b" if used, the script will automatically try to find the patient's T1s in a BIDS directory.
- "-t" for specifying whether to use the template more directly for the filling process, following the same workflow for bilateral lesions.
- "-E" to treat the lesion as an extra-axial pathology as described by Hou and colleagues in their recent work[4], by simply setting lesion voxels to 0.
- "-P" to follow a more pediatric friendly workflow, using the "NKI 10 and under" template and pediatric priors for tissue segmentation.
- "-F" if activated runs part 2 of VBG, including Freesurfer recon-all and subsequent steps, if not activated VBG simply generates the filled images and quits.
- One can also specify the location of the processing directory with the "-m" flag and similarly for the output with the "-o" flag. If one or neither is specified, the script falls back to a hardcoded intermediate and output directory.

#### 2. Supplementary results

The first SP parcellation attempt without using VBG lesion filling utilized a CFM like strategy, where the lesion was zero-filled using the extra-axial approach prior to recon-all (FreeSurferWiki, 2020a) (zF, N=44/56 completed/failed, grand average DSC = 0.230, standard deviation = 0.142). The second attempt used the images without any intervention beyond inserting an HD-BET (Isensee et al., 2019) brain mask into the recon-all (FreeSurferWiki, 2020a) workflow (non-VBG, N=48/52 completed/failed, grand average DSC = 0.675, and standard deviation = 0.281). Results from all completed parcellations are listed below in supplementary table 1 and supplementary figure 3.

**Supplementary table 1 – Unpaired DSC comparison results**

| **Unpaired DSC comparison results** | | | | |
|---|---|---|---|---|
| **Summary statistics** | | | | |
| **Groups** | Count | Average | | Std dev |
| **uVBG** | 100 | 0.889 | | 0.022 |
| **bVBG** | 25 | 0.901 | | 0.017 |
| **Non-VBG** | 48 | 0.676 | | 0.281 |
| **ANOVA: Single factor** | | | | |
| **Source of variation** | df | F | *P*-value | F critical |
| **Between groups** | 2 | 36.467 | <. 0001 | 3.049 |
| **Within groups** | 170 | | | |
| **Total** | 172 | | | |
| **Post hoc t-tests: Unpaired two-sample assuming unequal variances** | | | | |
| | df | t stat | *P* (two-tail) | t critical (two-tail) |
| **uVBG vs non-VBG** | 47 | 2.791 | < .01 | 2.012 |
| **bVBG vs non-VBG** | 48 | 3.079 | < .01 | 2.010 |

SP = synthetic patient, PAT = patient participant, SM = synthetic mass effect group, HCs = healthy control participants, HCs* = a-posterior warped HCs, uVBG = unilateral VBG, bVBG = bilateral VBG, DSC = dice similarity coefficient, uVBG* = uVBG corresponding to completed non-VBG parcellations, ANOVA = analysis of variance, df = degrees of freedom, Std dev = standard deviation, Stat = statistic

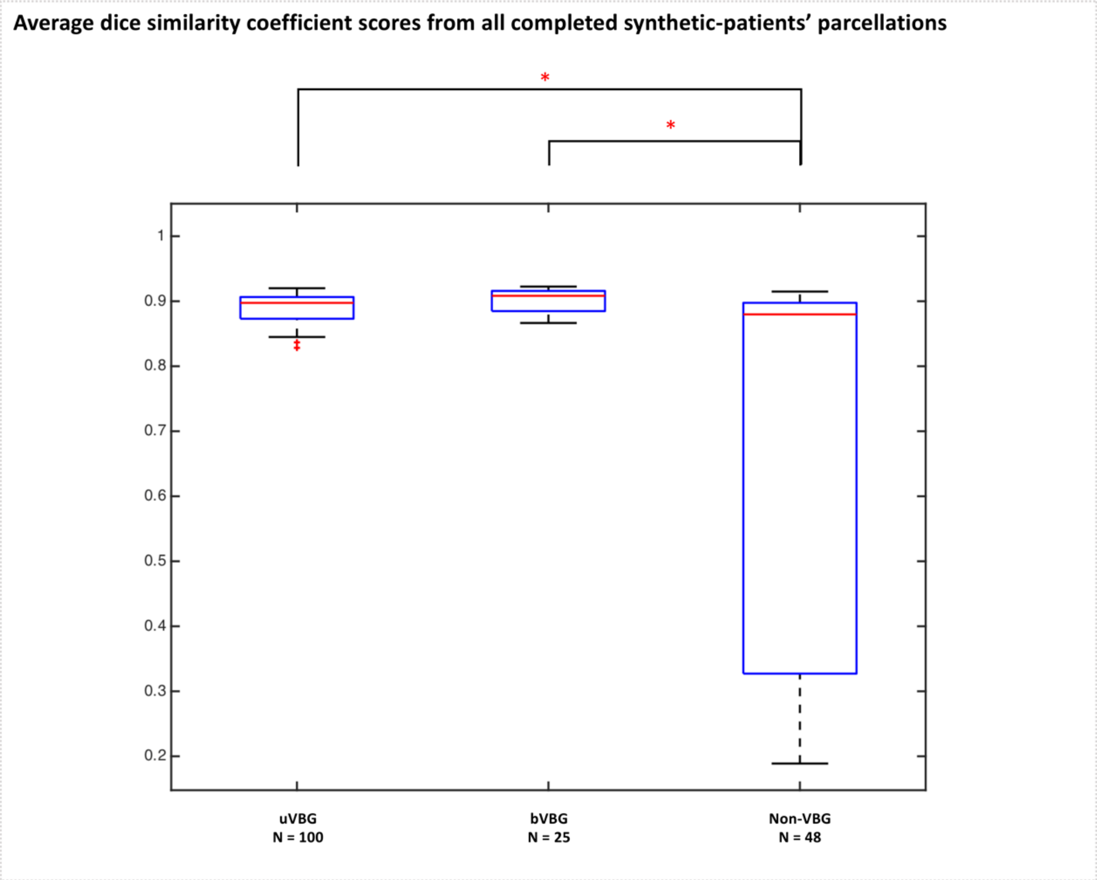


**Supplementary figure 3:** Average DSC values of all completed parcellations by each approach. Asterisks indicate statistically significant differences ($P < .01$)

##### 3. Supplementary figures referred to only in the main text:

- **Supplementary figure 4**

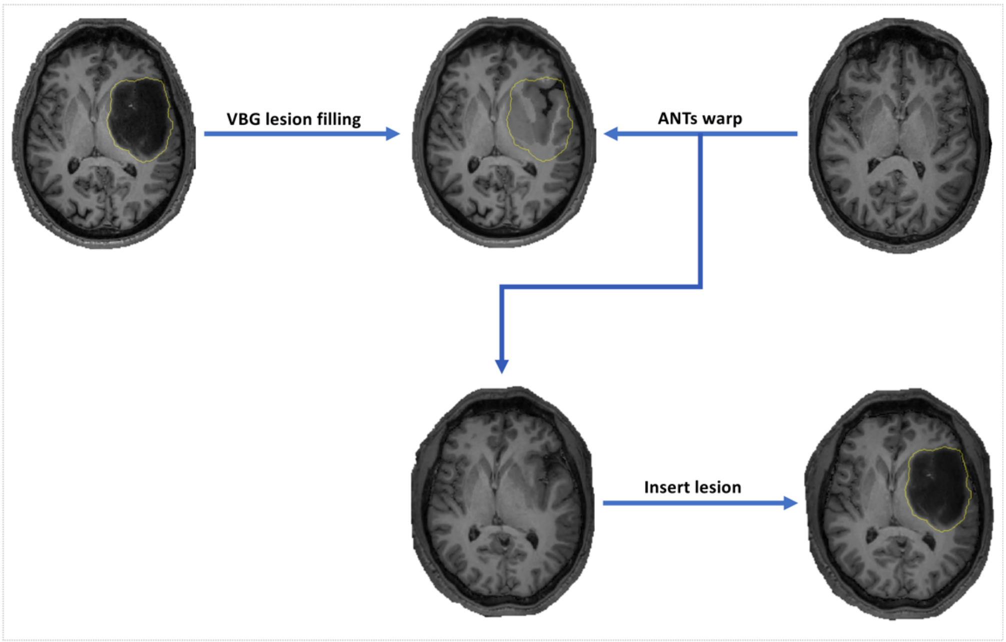


**Supplementary figure 4:** Axial slices showing generation of synthetic data from PAT and HC data. PAT = real patient participant, HC = heathy control, SM = synthetic mass effect group, SP = synthetic patient

- **Supplementary figure 5**

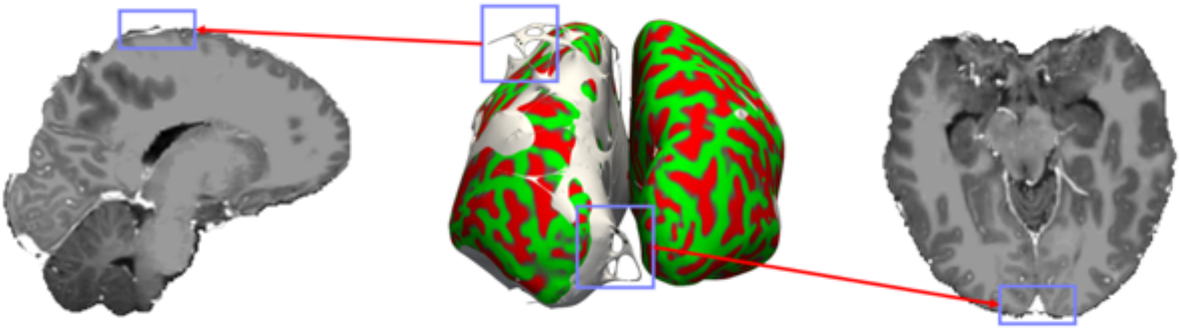

**Supplementary figure 5:** Surface topological errors in PT004, contrast enhanced T1 image after VBG filling. Middle image shows the inflated surface in white and the corrected surface overlaid in red/green. Violet rectangles indicate areas of error on the inflated uncorrected surface, shown in white and corresponding sagittal and axial slices.

- **Supplementary figure 6**

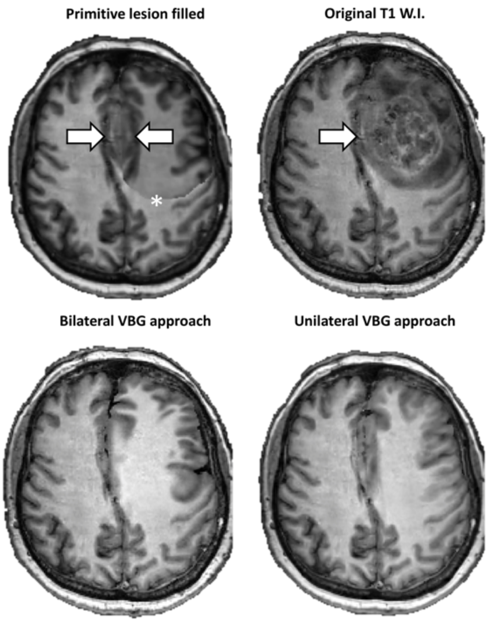


**Supplementary figure 6:** Top right shows input T1 WI with lesion, top left shows failed filling attempt. Note duplicated lesion component (arrows) and sharp interface (asterisk). uVBG and bVBG filled results are shown on the bottom.

- **Supplementary figure 7**

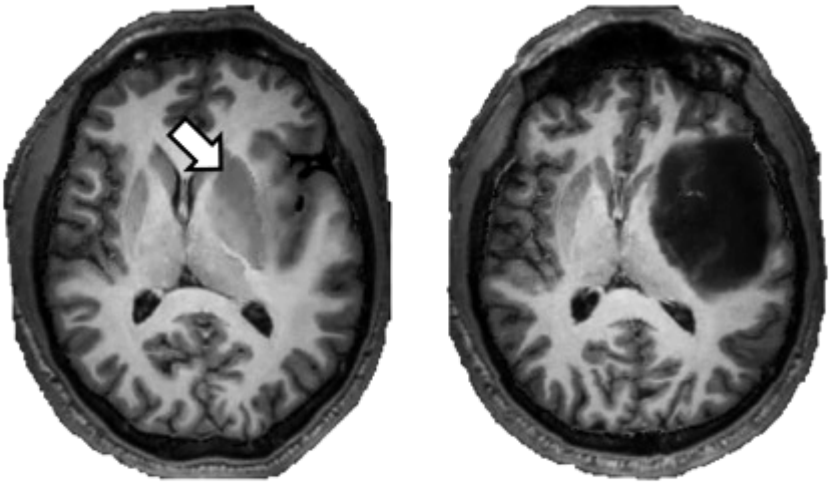

**Supplementary figure 7:** Example from a synthetic patient image, T1 with lesion is shown on the right, left shows the VBG filled brain, white arrow indicates an artificially enlarged left putamen and globus pallidus.
